# Prevalence and correlates of comorbid obstructive sleep apnea and restless legs syndrome in a population-based study from Benin, West Africa

**DOI:** 10.64898/2026.08.10.26360110

**Authors:** Ablo Prudence Wachinou, Abel Soumaho, Eudoscie Alitondé Djegbeton, Alima Koné, Hermionne Loko, Pervenche Mefo Fotso, Sandra Segoun, Denise Moussoro, Dieudonné Gnonlonfoun, Raphael Heinzer, Gildas Agodokpessi

## Abstract

**Purpose:** Comorbid restless legs syndrome (RLS) and obstructive sleep apnea (OSA) -(COROSA)- is poorly characterized in African populations. We estimated its prevalence and correlates in a population-based sample in Benin.

**Methods:** This was a cross-sectional analysis of 1,810 adults aged ≥25 years from the Benin Society and Sleep (BeSAS) study. RLS was defined by International RLS Study Group criteria and OSA by an apnea–hypopnea index ≥5 events/h on home respiratory polygraphy; COROSA required both. Correlates were assessed by multivariable logistic regression; multinomial models compared COROSA with OSA only, RLS only, and neither disorder. An exploratory analysis examined hypertension across eight mutually exclusive sleep-disorder groups, including comorbid insomnia, RLS and OSA (COMIROSA).

**Results:** COROSA prevalence was 3.5% (95% CI 2.7–4.4). Independent correlates were age 40–59 years (aOR 2.91, 95% CI 1.38–6.73), age ≥60 years (aOR 3.76, 1.61–9.35), rural residence (aOR 10.54, 4.94–25.43), overweight (aOR 2.29, 95% CI: 1.16–4.49), obesity (aOR 3.83, 95% CI: 1.75–8.31), and insomnia (aOR 2.49, 1.37–4.50). Relative to OSA only, COROSA was associated with hypertension and insomnia; relative to RLS only, obesity was the main distinguishing factor. Hypertension was associated with COROSA and COMIROSA, with a larger estimate for COMIROSA (aOR 4.03 vs 2.64).

**Conclusion:** COROSA affected 3.5% of adults in Benin and co-occurred with obesity, hypertension and insomnia. Screening for overlapping sleep disorders may improve identification of high-risk individuals. COMIROSA remains a hypothesis requiring validation in larger longitudinal studies.

**Brief summary:** The coexistence of obstructive sleep apnea and restless legs syndrome is insufficiently documented in African populations despite the growing burden of sleep disorders in the region. This study provides the first population-based study estimate of comorbid obstructive sleep apnea and restless legs syndrome in sub-Saharan Africa.

Our findings suggest that individuals with both disorders represent a clinically important subgroup with substantial cardiometabolic burden and insomnia. They also raise the possibility of an even more complex overlap involving chronic insomnia, restless legs syndrome, and obstructive sleep apnea, which warrants validation in future studies.

These results support more integrated approaches to screening and management of these coexisting sleep disorders in resource-limited settings.

## Introduction

Sleep disorders represent a growing public health concern worldwide, yet their burden remains insufficiently documented in many low- and middle-income countries [1–3], particularly in sub-Saharan Africa. Among these conditions, obstructive sleep apnea (OSA) and restless legs syndrome (RLS) stand out as two of the most common and clinically significant disorders, each associated with impaired sleep quality, reduced daytime functioning, and elevated cardiometabolic risk [4–6].

Although OSA and RLS have traditionally been studied as separate conditions, emerging evidence indicates that they frequently coexist within the same individual. This co-occurrence, hereafter referred to as COROSA (comorbid RLS and OSA) [6] or ComOSAR, [7] is increasingly recognized as a distinct clinical entity that may exacerbate sleep disruption and amplifies adverse health consequences beyond those associated with either disorder alone [5,7–10]. A recent systematic review estimated its prevalence to range from 10 to 36%, underscoring the need for systematic dual screening [6].

Despite the growing global interest in COROSA, data from African populations remain extremely scarce. The epidemiology of sleep disorders in the region is itself underexplored, and contextual factors such as urbanization, environmental exposures, and the dual burden of infectious or non-communicable diseases may influence both the occurrence and presentation of COROSA in ways not captured by studies conducted in high-income settings.

To address this knowledge gap, the present study aimed to (i) determine the prevalence of COROSA, (ii) identify its associated factors, and (iii) compare its clinical profile with isolated OSA and RLS in a population-based sample in Benin.

## Methods

### Study design and population

We conducted a population-based cross-sectional study between April 2018 and January 2020 in Benin, West Africa. Participants were recruited from the Benin Society and Sleep (BeSAS) study, which includes a rural (Tanve) and an urban (third sub-city of Cotonou) population. The study design and sampling procedures have been described previously [11,12].

In brief, rural participants were drawn from the Tanve Health Study (TAHES), a longitudinal cohort study investigating cardiovascular diseases and their risk factors in Benin. All consenting individuals enrolled in this cohort were included in the BeSAS study [13]. Urban participants were randomly selected from nine of the thirteen districts comprising the third sub-city of Cotonou. Households were chosen using a proportional sampling technique, and within each selected household, all eligible and consenting individuals were recruited [11,12].

### Inclusion criteria and sample size

Participants aged ≥ 25 years enrolled in the BeSAS study who had valid overnight respiratory polygraphy recordings were included. No formal sample size calculation was performed, as the study was based on an existing population cohort.

### Data collection

#### Main outcome

The main outcome was comorbid restless legs syndrome (RLS) and obstructive sleep apnea (OSA), defined as COROSA. RLS and OSA were assessed using the standardized International Restless Legs Questionnaire and overnight respiratory polygraphy, respectively.

RLS was diagnosed based on the four mandatory criteria established by the International Restless Legs Syndrome Study Group (IRLSSG), published in 1995 and revised in 2003 by the US National Institutes of Health (NIH) [14]:

- Imperative need to move the lower limbs, often associated with uncomfortable, unpleasant sensations. The upper limbs and other parts of the body are less affected;
- Appearance or aggravation of symptoms during periods of rest or inactivity, particularly when lying down or sitting;
- Relief or emission of symptoms on exertion, at least temporarily, as long as the activity lasts;
- Appearance or marked aggravation of symptoms in the evening or at night.

Respiratory polygraphy was performed using ApneaLink Plus devices (Resmed, Bella Vista, Australia), a type-3 portable recorder measuring airflow via a nasal pressure sensor, respiratory effort (thoracic movement), and pulse oximetry (Nonin; Nonin Medical, Plymouth, MA, USA). Devices were installed at participants’ homes between 20:00 and 22:00 by four trained research assistants and collected the following morning. Participants were asked to report lights-off and lights-on times. Polygraphy data were converted to European Data Format files and analyzed using Noxturnal software (version 6.2; Nox Medical, Reykjavik, Iceland). Recordings were manually scored by a certified sleep physician (APW) with over 10 years of experience in respiratory sleep medicine, blinded to the participants’ clinical data. Only recordings with at least 4 hours of valid airflow, respiratory effort, and pulse oximetry signals were included in the analysis. Respiratory events were scored according to the 2012 American Academy of Sleep Medicine manual [15]. Sleep apnea was defined as a ≥ 90% reduction in airflow lasting at least 10 seconds, while hypopnea was defined as a ≥ 30% reduction in airflow for at least 10 seconds associated with a ≥ 3% oxygen desaturation. The apnea–hypopnea index (AHI) was calculated as the total number of apneas and hypopneas divided by the estimated sleep time reported by participants. OSA was defined as an AHI ≥ 5 events/hour.

### Covariates

Sociodemographic characteristics (age, sex and area of residence) and behavioral factors (alcohol consumption, tobacco use, and physical activity) were collected through face-to-face interviews.

Anthropometric measurements were obtained using standard protocols. Height was measured with a rigid measuring device, and weight using a calibrated mechanical device (Seca, Hamburg, Germany), with participants standing barefoot and wearing light clothing. Body mass index (BMI) was calculated as weight divided by height squared. BMI was categorized as: < 18.5 kg/m² (underweight), 18.5 to < 25 kg/m² (normal weight), 25 to < 30 kg/m² (overweight), and ≥ 30 kg/m² (obese) [16].

Blood pressure was measured three times on both arms (Spengler, France), with the mean of the second and third readings calculated for each arm. Hypertension was defined as systolic blood pressure ≥ 140 mmHg or diastolic blood pressure ≥ 90 mmHg on either arm, or self-reported antihypertensive treatment [16].

Fasting blood glucose was measured using a glucometer (Accuchek Performa^®^, Roche Diagnostics, Basel, Switzerland). Diabetes mellitus was defined as fasting blood glucose levels ≥ 7 mmol/L or a self-reported medical history of diabetes mellitus. Sleep quality was assessed using the Pittsburgh Sleep Quality Index (PSQI), which evaluates the subjective sleep patterns over a 1-month time period. The total score ranges from 0 to 21, and a score ≥ 6 suggests poor sleep quality [17].

Insomnia severity was evaluated using the Insomnia Severity Index (ISI), with a total score ranging from 0 to 28. A score > 7 indicates the presence of insomnia [18].

Daytime sleepiness was assessed using the Epworth Sleepiness Scale (ESS), ranging from 0 to 24, with a score ≥ 11 indicating excessive daytime sleepiness [19].

### Data analysis

Categorical variables were summarized as frequencies and percentages. Group comparisons were performed using Pearson Chi-squared test or Fisher’s exact test, as appropriate.

### Main analysis

The primary analysis assessed factors associated with COROSA using binary logistic regression. Variables associated with COROSA at p-value < 0.25 in the univariable analyses were included in the multivariable model. Adjusted odds ratios (aORs) and their 95% confidence intervals (CIs) were reported.

Multicollinearity among covariates was assessed using variance inflation factors (VIF), with values <5 considered acceptable. Model performance was evaluated in terms of calibration and discrimination using the Hosmer–Lemeshow goodness-of-fit test and the area under the receiver operating characteristic curve (AUC), respectively.

### Secondary analyses

To examine the factors associated with different sleep-disorder phenotypes, participants were initially classified into four mutually exclusive groups: no disorder, OSA only, RLS only, and COROSA. A multinomial logistic regression model was fitted using the no-disorder group as the reference category. A backward stepwise selection procedure was used to derive the final model, retaining statistically significant variables. Model performance was assessed using overall classification accuracy and the mean and/or class-specific AUC values.

To further characterize the clinical profile of COROSA, additional multinomial logistic regression analyses were conducted using the OSA-only and RLS-only groups as alternative reference categories. These analyses enabled direct comparisons between participants with COROSA and those with either isolated OSA or isolated RLS. The covariates included in the primary multivariable model were retained, and aORs with 95% CIs were estimated to identify factors distinguishing COROSA from each isolated sleep disorder.

Finally, as an exploratory, hypothesis-generating analysis, we examined whether the addition of insomnia to the RLS-OSA comorbidity pattern defined a distinct phenotype – comorbid insomnia, restless legs syndrome, and obstructive sleep apnea (COMIROSA) with a potentially different cardiometabolic risk profile. This analysis was motivated by evidence that RLS and OSA frequently coexist and their coexistence is accompanied by a greater burden of insomnia symptoms [20] and by their potentially cumulative effects on sleep fragmentation, autonomic activation, and cardiometabolic risk. Participants were then reclassified into eight mutually exclusive phenotypic groups: no sleep disorder, OSA only, RLS only, insomnia only, comorbid insomnia and OSA (COMISA), comorbid RLS and insomnia, COROSA, and COMIROSA. Using the no-disorder group as the reference category, multinomial logistic regression was used to estimate the adjusted associations between each phenotype and hypertension. Models were adjusted for age, sex, area of residence, body mass index, diabetes status, and sleep quality as assessed by the Pittsburgh Sleep Quality Index. Particular attention was given to whether the association with hypertension was stronger for COMIROSA than for COROSA. Odds ratios for all phenotypes were displayed in a forest plot. To test our hypothesis, that adding insomnia may intensify the cardiovascular risk of the RLS-OSA overlap, we directly compared the hypertension coefficients of the COMIROSA and COROSA phenotypes using a Wald test.

### Sensitivity analyses

To evaluate the robustness of our findings, the main and secondary analyses were repeated using two more stringent ISI thresholds (≥ 10 and ≥ 15) instead of the predefined threshold (> 7) to define insomnia.

Statistical significance was set at p-value < 0.05. All analyses were performed using R software version 4.4.2 (R Foundation for Statistical Computing, Vienna, Austria).

## Results

### Characteristics of the study population

A total of 1,810 participants were included in this study, with a mean age of 45.5 ± 14.8 years. The majority were younger than 60 years (81.2%), predominantly female (64.2%), and almost evenly distributed between rural (52.8%) and urban (47.2%) areas (Table 1).

**Table 1:**
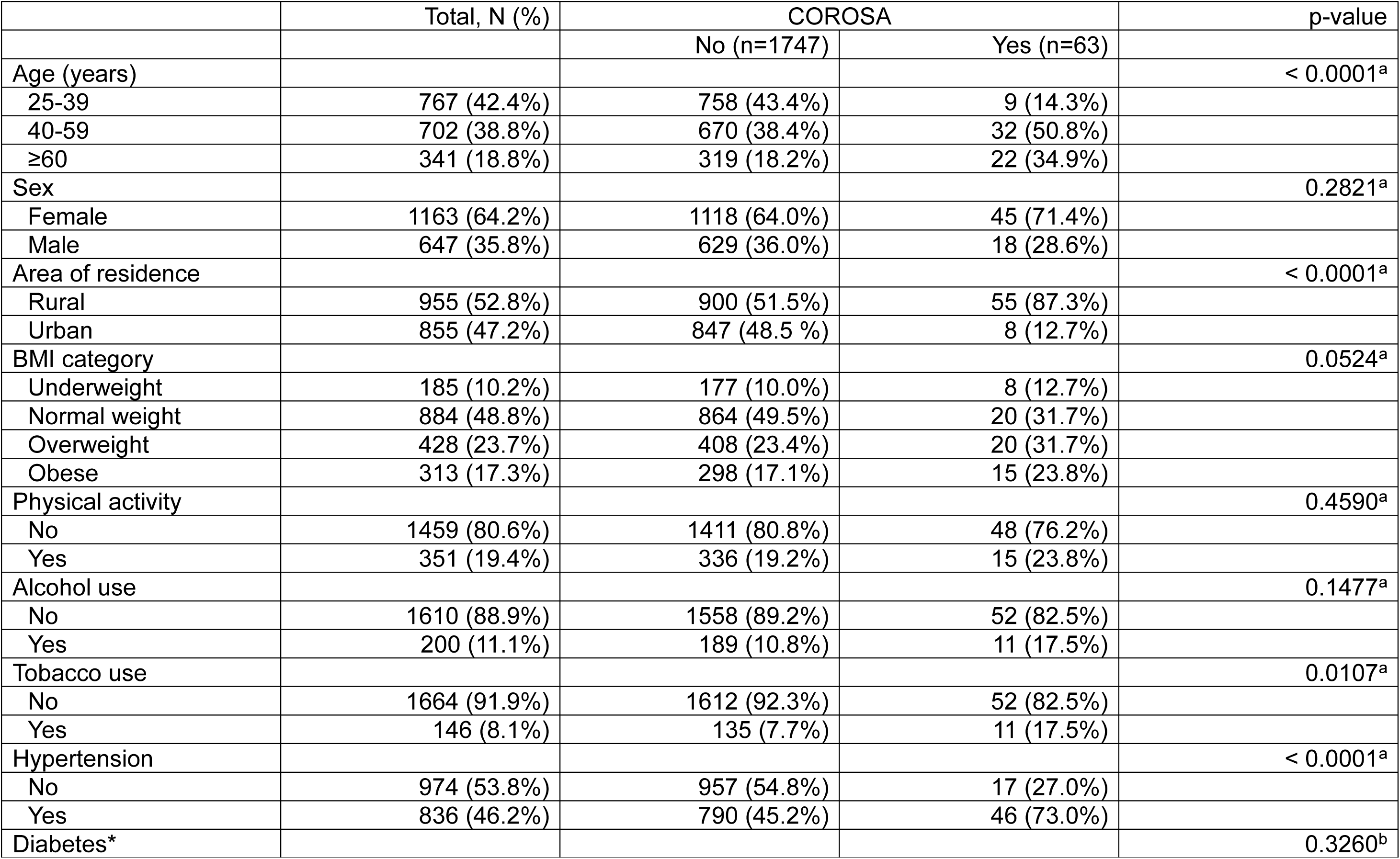
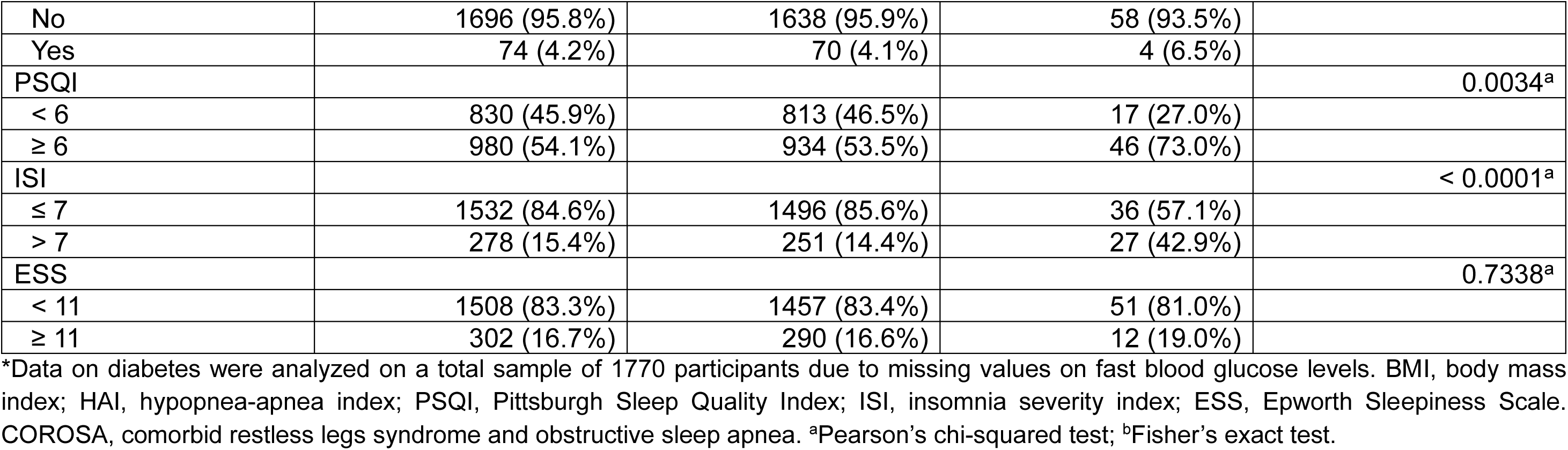
Baseline characteristics of BeSAS participants according to the COROSA status (N=1810)

|  | Total, N (%) | COROSA |  | p-value |
| --- | --- | --- | --- | --- |
|  |  | No (n=1747) | Yes (n=63) |  |
| Age (years) |  |  |  | < 0.0001<sup>a</sup> |
| 25-39 | 767 (42.4%) | 758 (43.4%) | 9 (14.3%) |  |
| 40-59 | 702 (38.8%) | 670 (38.4%) | 32 (50.8%) |  |
| ≥60 | 341 (18.8%) | 319 (18.2%) | 22 (34.9%) |  |
| Sex |  |  |  | 0.2821<sup>a</sup> |
| Female | 1163 (64.2%) | 1118 (64.0%) | 45 (71.4%) |  |
| Male | 647 (35.8%) | 629 (36.0%) | 18 (28.6%) |  |
| Area of residence |  |  |  | < 0.0001<sup>a</sup> |
| Rural | 955 (52.8%) | 900 (51.5%) | 55 (87.3%) |  |
| Urban | 855 (47.2%) | 847 (48.5 %) | 8 (12.7%) |  |
| BMI category |  |  |  | 0.0524<sup>a</sup> |
| Underweight | 185 (10.2%) | 177 (10.0%) | 8 (12.7%) |  |
| Normal weight | 884 (48.8%) | 864 (49.5%) | 20 (31.7%) |  |
| Overweight | 428 (23.7%) | 408 (23.4%) | 20 (31.7%) |  |
| Obese | 313 (17.3%) | 298 (17.1%) | 15 (23.8%) |  |
| Physical activity |  |  |  | 0.4590<sup>a</sup> |
| No | 1459 (80.6%) | 1411 (80.8%) | 48 (76.2%) |  |
| Yes | 351 (19.4%) | 336 (19.2%) | 15 (23.8%) |  |
| Alcohol use |  |  |  | 0.1477<sup>a</sup> |
| No | 1610 (88.9%) | 1558 (89.2%) | 52 (82.5%) |  |
| Yes | 200 (11.1%) | 189 (10.8%) | 11 (17.5%) |  |
| Tobacco use |  |  |  | 0.0107<sup>a</sup> |
| No | 1664 (91.9%) | 1612 (92.3%) | 52 (82.5%) |  |
| Yes | 146 (8.1%) | 135 (7.7%) | 11 (17.5%) |  |
| Hypertension |  |  |  | < 0.0001<sup>a</sup> |
| No | 974 (53.8%) | 957 (54.8%) | 17 (27.0%) |  |
| Yes | 836 (46.2%) | 790 (45.2%) | 46 (73.0%) |  |
| Diabetes* |  |  |  | 0.3260<sup>b</sup> |
| No | 1696 (95.8%) | 1638 (95.9%) | 58 (93.5%) |  |
| Yes | 74 (4.2%) | 70 (4.1%) | 4 (6.5%) |  |
| PSQI |  |  |  | 0.0034<sup>a</sup> |
| < 6 | 830 (45.9%) | 813 (46.5%) | 17 (27.0%) |  |
| ≥ 6 | 980 (54.1%) | 934 (53.5%) | 46 (73.0%) |  |
| ISI |  |  |  | < 0.0001<sup>a</sup> |
| ≤ 7 | 1532 (84.6%) | 1496 (85.6%) | 36 (57.1%) |  |
| > 7 | 278 (15.4%) | 251 (14.4%) | 27 (42.9%) |  |
| ESS |  |  |  | 0.7338<sup>a</sup> |
| < 11 | 1508 (83.3%) | 1457 (83.4%) | 51 (81.0%) |  |
| ≥ 11 | 302 (16.7%) | 290 (16.6%) | 12 (19.0%) |  |

\*Data on diabetes were analyzed on a total sample of 1770 participants due to missing values on fast blood glucose levels. BMI, body mass index; HAI, hypopnea-apnea index; PSQI, Pittsburgh Sleep Quality Index; ISI, insomnia severity index; ESS, Epworth Sleepiness Scale. COROSA, comorbid restless legs syndrome and obstructive sleep apnea. <sup>a</sup>Pearson's chi-squared test; <sup>b</sup>Fisher's exact test.

Nearly half of participants had a normal BMI (48.8%). Hypertension, diabetes, poor sleep quality, insomnia, and excessive daytime sleepiness were present in 46.2%, 4.2%, 54.1%, 15.4% and 16.7% of participants, respectively.

### Prevalence of COROSA

The prevalence of COROSA was 3.5% (63/1810; 95% CI: 2.7 – 4.4%). Participants with COROSA were significantly older, more likely to reside in rural areas, to be overweight or obese, smokers, hypertensive, poor sleepers, and to report insomnia symptoms (Table 1).

### Factors associated with COROSA

After adjustment in a binary logistic regression model, participants aged 40 years and above were significantly more likely to have COROSA compared to those aged 25 to 39 years (40-59 years: aOR 2.91, 95% CI: 1.38–6.73; ≥60 years: aOR 3.76, 95% CI: 1.61–9.35). Rural residence was independently associated with COROSA (aOR 10.54, 95% CI: 4.94–25.42) compared with urban residents.

In addition, overweight (aOR 2.29, 95% CI: 1.16–4.49), obesity (aOR 3.83, 95% CI: 1.75–8.31), hypertension (aOR 2.26, 95% CI: 1.25–4.27), and insomnia (aOR 2.49, 95% CI: 1.37–4.50) were independently associated with increased odds of COROSA (Table 2).

**Table 2:** Factors associated with COROSA among BeSAS participants – binary logistic regression analysis (N=1810)

|  | Univariable model |  |  | Multivariable model |  |  |
| --- | --- | --- | --- | --- | --- | --- |
|  | cOR | 95% CI | p-value | aOR | 95% CI | p-value |
| Age (years) |  |  |  |  |  |  |
| 25-39 | 1 |  |  | 1 |  |  |
| 40-59 | 4.02 | 1.98 – 9.01 | 0.0003 | 2.91 | 1.38 – 6.73 | 0.0075 |
| ≥60 | 5.81 | 2.73 – 13.43 | < 0.0001 | 3.76 | 1.61 – 9.35 | 0.0028 |
| Sex |  |  |  |  |  |  |
| Female | 1 |  |  | 1 |  |  |
| Male | 0.71 | 0.40 – 1.22 | 0.2286 | 0.90 | 0.46 – 1.70 | 0.7523 |
| Area of residence |  |  |  |  |  |  |
| Urban | 1 |  |  | 1 |  |  |
| Rural | 6.47 | 3.25 – 14.77 | < 0.0001 | 10.54 | 4.94 – 25.42 | < 0.0001 |
| BMI category |  |  |  |  |  |  |
| Normal weight | 1 |  |  | 1 |  |  |
| Underweight | 1.95 | 0.79 – 4.35 | 0.1166 | 1.30 | 0.51 – 3.01 | 0.5569 |
| Overweight | 2.12 | 1.12 – 4.00 | 0.0198 | 2.29 | 1.16 – 4.49 | 0.0156 |
| Obese | 2.17 | 1.08 – 4.28 | 0.0256 | 3.83 | 1.75 – 8.31 | < 0.0001 |
| Physical activity |  |  |  |  |  |  |
| No | 1 |  |  | - |  |  |
| Yes | 1.31 | 0.70 – 2.32 | 0.3681 | - | - | - |
| Alcohol use |  |  |  |  |  |  |
| No | 1 |  |  | 1 |  |  |
| Yes | 1.74 | 0.85 – 3.27 | 0.1026 | 1.35 | 0.59 – 2.89 | 0.4522 |
| Tobacco use |  |  |  |  |  |  |
| No | 1 |  |  | 1 |  |  |
| Yes | 2.53 | 1.23 – 4.78 | 0.0070 | 1.33 | 0.58 – 2.89 | 0.4799 |
| Hypertension |  |  |  |  |  |  |
| No | 1 |  |  | 1 |  |  |
| Yes | 3.28 | 1.90 – 5.92 | < 0.0001 | 2.26 | 1.25 – 4.27 | 0.0089 |
| Diabetes* |  |  |  | - |  |  |
| No | 1 |  |  | - |  |  |
| Yes | 1.61 | 0.48 – 4.07 | 0.3675 | - | - | - |
| PSQI |  |  |  |  |  |  |
| < 6 | 1 |  |  | 1 |  |  |
| ≥ 6 | 2.36 | 1.37 – 4.25 | 0.0029 | 1.17 | 0.63 – 2.26 | 0.6214 |
| ISI |  |  |  |  |  |  |
| ≤ 7 | 1 |  |  | 1 |  |  |
| > 7 | 4.47 | 2.65 – 7.47 | < 0.0001 | 2.49 | 1.37 – 4.50 | 0.0025 |
| ESS |  |  |  |  |  |  |
| < 11 | 1 |  |  | - |  |  |
| ≥ 11 | 1.18 | 0.59 – 2.17 | 0.6091 | - | - | - |

\*Univariable analysis on diabetes was analyzed on a total sample of 1770 participants due to missing values on fast blood glucose levels. The multivariable model was adjusted for age, sex, area of residence, BMI category, alcohol use, tobacco use, hypertension, PSQI and insomnia severity. cOR, crude odds-ratio; aOR, adjusted odds-ratio; CI, confidence interval; BMI, body mass index; PSQI, Pittsburgh Sleep Quality Index; ISI, insomnia severity index. Model performance – Hosmer-Lemeshow Goodness-of-Fit Test: p-value = 0.938; AUC: 0.841 (95% CI: 0.791 – 0.892)

### Comparative analysis of determinants of OSA-only, RLS-only and COROSA

Distribution of participants characteristics per sleep disorders phenotype (None, OSA-only, RLS-only, COROSA) is shown in supplement Table S1.

To compare determinants of COROSA with those of each isolated sleep disorder, we first performed an univariable multinomial logistic regression analyses taking the “none” group as reference (Table 3). Older age was associated with all three outcomes, with the highest odds observed for RLS-only and COROSA. Male sex, overweight/obesity, alcohol consumption, tobacco use, hypertension, diabetes, and insomnia were associated with increased odds of OSA-only, whereas rural residence, tobacco use, hypertension, poor sleep quality, and insomnia were significantly associated with increased odds of RLS-only and COROSA.

**Table 3:** Univariable analyses of factors associated with COROSA among BeSAS participants – multinomial logistic regression (N=1810)

|  | OSA only |  | RLS only |  | COROSA |  |
| --- | --- | --- | --- | --- | --- | --- |
|  | cOR (95% CI) | p-value | cOR (95% CI) | p-value | cOR (95% CI) | p-value |
| Age (years) |  |  |  |  |  |  |
| 25-39 | 1 |  | 1 |  | 1 |  |
| 40-59 | 2.73 (2.19 – 3.41) | < 0.0001 | 3.80 (1.17 – 8.45) | 0.0001 | 6.10 (2.87 – 12.94) | < 0.0001 |
| ≥60 | 3.50 (2.65 – 4.62) | < 0.0001 | 8.96 (3.96 – 20.26) | < 0.0001 | 10.39 (4.67 – 23.13) | < 0.0001 |
| Sex |  |  |  |  |  |  |
| Female | 1 |  | 1 |  | 1 |  |
| Male | 1.75 (1.43 – 2.14) | < 0.0001 | 0.84 (0.44 – 1.60) | 0.5885 | 0.90 (0.51 – 1.58) | 0.7107 |
| Area of residence |  |  |  |  |  |  |
| Urban | 1 |  | 1 |  | 1 |  |
| Rural | 0.52 (0.43 – 0.63) | < 0.0001 | 3.70 (1.71 – 7.98) | 0.0008 | 5.08 (2.30 – 10.78) | < 0.0001 |
| BMI category |  |  |  |  |  |  |
| Underweight/Normal weight | 1 |  | 1 |  | 1 |  |
| Overweight/Obese | 2.70 (2.21 – 3.30) | < 0.0001 | 0.93 (0.49 – 1.76) | 0.8203 | 2.82 (1.69 – 4.73) | < 0.0001 |
| Physical activity |  |  |  |  |  |  |
| No | 1 |  | 1 |  | 1 |  |
| Yes | 1.17 (0.92 – 1.49) | 0.2021 | 1.19 (0.58 – 2.43) | 0.6382 | 1.41 (0.77 – 2.57) | 0.2635 |
| Alcohol use |  |  |  |  |  |  |
| No | 1 |  | 1 |  | 1 |  |
| Yes | 1.67 (1.22 – 2.27) | 0.0011 | 1.51 (0.62 – 3.64) | 0.3639 | 2.23 (1.12 – 4.43) | 0.0222 |
| Tobacco use |  |  |  |  |  |  |
| No | 1 |  | 1 |  | 1 |  |
| Yes | 1.48 (1.03 – 2.13) | 0.0362 | 3.97 (1.89 – 8.34) | 0.0003 | 3.19 (1.58 – 6.43) | 0.0011 |
| Hypertension |  |  |  |  |  |  |
| No | 1 |  | 1 |  | 1 |  |
| Yes | 2.08 (1.71 – 2.53) | < 0.0001 | 1.82 (1.02 – 3.25) | 0.0435 | 4.53 (2.56 – 8.01) | < 0.0001 |
| Diabetes* |  |  |  |  |  |  |
| No | 1 |  | 1 |  | 1 |  |
| Yes | 2.59 (1.57 – 4.27) | < 0.0001 | 0.81 (0.11 – 6.08) | 0.8342 | 2.61 (0.88 – 7.76) | 0.0837 |
| PSQI |  |  |  |  |  |  |
| < 6 | 1 |  | 1 |  | 1 |  |
| ≥ 6 | 1.07 (0.88 – 1.30) | 0.4764 | 2.01 (1.08 – 3.74) | 0.0284 | 2.47 (1.40 – 4.37) | 0.0018 |
| ISI |  |  |  |  |  |  |
| ≤ 7 | 1 |  | 1 |  | 1 |  |
| > 7 | 1.44 (1.09 – 1.89) | 0.0098 | 3.33 (1.75 – 6.31) | 0.0002 | 5.48 (3.21 – 9.36) | < 0.0001 |
| ESS |  |  |  |  |  |  |
| < 11 | 1 |  | 1 |  | 1 |  |
| ≥ 11 | 1.24 (0.96 – 1.60) | 0.1057 | 1.28 (0.61 – 2.69) | 0.5195 | 1.30 (0.68 – 2.50) | 0.4247 |

\*Data on diabetes were analyzed on a total sample of 1770 participants due to missing values on fast blood glucose levels. BMI, body mass index; PSQI, Pittsburgh Sleep Quality Index; ISI, insomnia severity index; OSA, obstructive sleep apnea; RLS, restless legs syndrome; COROSA, comorbid restless legs syndrome and obstructive sleep apnea; cOR, crude odds-ratio; CI, confidence interval.

In the multivariable analyses, (Table 4a), older age was associated with all three outcomes, but the associations appeared stronger for RLS-only and COROSA than for OSA-only. Male sex and overweight/obesity were mainly associated with OSA only, while rural residence and higher ISI scores showed strong associations for RLS-only and COROSA. Hypertension was also associated with OSA-only and COROSA, with higher estimates for COROSA.

**Table 4a:** Multivariable analyses of factors associated with COROSA among BeSAS participants – multinomial logistic regression (N=1770)

|  | OSA only |  | RLS only |  | COROSA |  |
| --- | --- | --- | --- | --- | --- | --- |
|  | aOR (95% CI) | p-value | aOR (95% CI) | p-value | aOR (95% CI) | p-value |
| Age (years) |  |  |  |  |  |  |
| 25-39 | 1 |  | 1 |  | 1 |  |
| 40-59 | 2.28 (1.79 – 2.90) | < 0.0001 | 3.61 (1.59 – 8.22) | 0.0022 | 4.28 (1.94 – 9.41) | 0.0003 |
| ≥60 | 3.14 (2.30 – 4.28) | < 0.0001 | 7.89 (3.30 – 18.8) | < 0.0001 | 7.21 (3.05 – 17.1) | < 0.0001 |
| Sex |  |  |  |  |  |  |
| Female | 1 |  | 1 |  | 1 |  |
| Male | 2.40 (1.91 – 3.03) | < 0.0001 | 0.95 (0.48 – 1.87) | 0.8723 | 1.55 (0.84 – 2.87) | 0.1610 |
| Area of residence |  |  |  |  |  |  |
| Urban | 1 |  | 1 |  | 1 |  |
| Rural | 0.78 (0.63 – 0.97) | 0.0286 | 4.16 (1.87 – 9.27) | 0.0004 | 9.96 (4.35 – 22.8) | < 0.0001 |
| BMI category |  |  |  |  |  |  |
| Underweight/Normal weight | 1 |  | 1 |  | 1 |  |
| Overweight/Obese | 2.89 (2.28 – 3.67) | < 0.0001 | 1.31 (0.65 – 2.61) | 0.4489 | 4.17 (2.33 – 7.49) | < 0.0001 |
| Hypertension |  |  |  |  |  |  |
| No | 1 |  | 1 |  | 1 |  |
| Yes | 1.27 (1.01 – 1.60) | 0.0398 | 1.10 (0.58 – 2.08) | 0.7745 | 2.48 (1.33 – 4.62) | 0.0043 |
| ISI |  |  |  |  |  |  |
| ≤ 7 | 1 |  | 1 |  | 1 |  |
| > 7 | 1.21 (0.90 – 1.64) | 0.2042 | 2.29 (1.17 – 4.50) | 0.0157 | 3.18 (1.78 – 5.68) | < 0.0001 |

The multinomial logistic regression analysis was performed on 1770 participants due to missing values on fast blood glucose levels. During the backward stepwise selection procedure, the following variables were not retained in the final model because they were not statistically significant: physical activity, alcohol use, tobacco use, PSQI, and diabetes. “No OSA and no RLS” was used as reference for the analysis. BMI, body mass index; ISI, insomnia severity index; OSA, obstructive sleep apnea; RLS, restless legs syndrome. COROSA, comorbid restless legs syndrome and obstructive sleep apnea; OR, odds-ratio; aOR, adjusted odds-ratio; CI, confidence interval. Model accuracy: 0.6378. AUC per class: 0.723 (None); 0.710 (OSA only); 0.754 (RLS only); 0.838 (COROSA). Mean AUC: 0.756.

To clarify which of the features specifically differentiated COROSA from each single disorder, the analysis was then repeated using OSA-only and RLS-only as reference categories. Compared to isolated OSA (Table 4b), COROSA was marked by higher odds of hypertension and insomnia symptoms, alongside rural residence. By contrast, relative to isolated RLS (Table 4c), overweight/obesity was the only factor independently associated with COROSA; the association with hypertension was of similar magnitude to that observed against OSA-only but did not reach statistical significance (p = 0.0612), and insomnia symptoms no longer distinguished the two groups (p = 0.4375).

**Table 4b:** Multivariable analyses of factors associated with COROSA among BeSAS participants – multinomial logistic regression using OSA-only as the reference category (N=1770)

|  | None |  | RLS only |  | COROSA |  |
| --- | --- | --- | --- | --- | --- | --- |
|  | aOR (95% CI) | p-value | aOR (95% CI) | p-value | aOR (95% CI) | p-value |
| **Age (years)** |  |  |  |  |  |  |
| 25-39 | 1 |  | 1 |  | 1 |  |
| 40-59 | 0.44 (0.34 – 0.56) | < 0.0001 | 1.58 (0.69 – 3.64) | 0.2805 | 1.88 (0.85 – 4.15) | 0.1213 |
| ≥60 | 0.32 (0.23 – 0.43) | < 0.0001 | 2.51 (1.04 – 6.06) | 0.0403 | 2.30 (0.97 – 5.45) | 0.0589 |
| **Sex** |  |  |  |  |  |  |
| Female | 1 |  | 1 |  | 1 |  |
| Male | 0.42 (0.33 – 0.52) | < 0.0001 | 0.39 (0.20 – 0.78) | 0.0076 | 0.65 (0.35 – 1.19) | 0.1614 |
| **Area of residence** |  |  |  |  |  |  |
| Urban | 1 |  | 1 |  | 1 |  |
| Rural | 1.28 (1.03 – 1.59) | 0.0286 | 5.32 (2.38 – 11.87) | < 0.0001 | 12.7 (5.58 – 29.03) | < 0.0001 |
| **BMI category** |  |  |  |  |  |  |
| Underweight/Normal weight | 1 |  | 1 |  | 1 |  |
| Overweight/Obese | 0.35 (0.27 – 0.44) | < 0.0001 | 0.45 (0.23 – 0.90) | 0.0249 | 1.44 (0.80 – 2.58) | 0.2186 |
| **Hypertension** |  |  |  |  |  |  |
| No | 1 |  | 1 |  | 1 |  |
| Yes | 0.79 (0.63 – 0.99) | 0.0399 | 0.86 (0.45 – 1.64) | 0.6567 | **1.95 (1.04 – 3.65)** | **0.0366** |
| **ISI** |  |  |  |  |  |  |
| ≤ 7 | 1 |  | 1 |  | 1 |  |
| > 7 | 0.82 (0.61 – 1.11) | 0.2042 | 1.89 (0.96 – 3.72) | 0.0659 | **2.62 (1.47 – 4.66)** | **0.0011** |

The multinomial logistic regression analysis was performed on 1770 participants due to missing values on fast blood glucose levels. During the backward stepwise selection procedure, the following variables were not retained in the final model because they were not statistically significant: physical activity, alcohol use, tobacco use, PSQI, and diabetes. OSA-only was used as reference for the analysis. BMI, body mass index; ISI, insomnia severity index; OSA, obstructive sleep apnea; RLS, restless legs syndrome. COROSA, comorbid restless legs syndrome and obstructive sleep apnea; OR, odds-ratio; aOR, adjusted odds-ratio; CI, confidence interval. Model accuracy: 0.6378. Mean AUC: 0.756. AUC per class: 0.723 (None); 0.710 (OSA only); 0.754 (RLS only); 0.838 (COROSA). Mean AUC: 0.756.

**Table 4c:** Multivariable analyses of factors associated with COROSA among BeSAS participants – multinomial logistic regression using RLS-only as the reference category.

|  | None |  | OSA only |  | COROSA |  |
| --- | --- | --- | --- | --- | --- | --- |
|  | aOR (95% CI) | p-value | aOR (95% CI) | p-value | aOR (95% CI) | p-value |
| Age (years) |  |  |  |  |  |  |
| 25-39 | 1 |  | 1 |  | 1 |  |
| 40-59 | 0.28 (0.12 – 0.63) | 0.0022 | 0.63 (0.27 – 1.45) | 0.2803 | 1.18 (0.39 – 3.61) | 0.7656 |
| ≥60 | 0.13 (0.05 – 0.30) | < 0.0001 | 0.40 (0.16 – 0.96) | 0.0402 | 0.91 (0.28 – 2.98) | 0.8821 |
| Sex |  |  |  |  |  |  |
| Female | 1 |  | 1 |  | 1 |  |
| Male | 1.06 (0.54 – 2.09) | 0.8723 | 2.54 (1.28 – 5.04) | 0.0076 | 1.64 (0.68 – 3.94) | 0.2671 |
| Area of residence |  |  |  |  |  |  |
| Urban | 1 |  | 1 |  | 1 |  |
| Rural | 0.24 (0.11 – 0.54) | 0.0005 | 0.19 (0.08 – 0.42) | < 0.0001 | 2.39 (0.78 – 7.37) | 0.1285 |
| **BMI category** |  |  |  |  |  |  |
| **Underweight/Normal weight** | 1 |  | 1 |  | **1** |  |
| **Overweight/Obese** | 0.77 (0.38 – 1.53) | 0.4489 | 2.21 (1.11 – 4.44) | 0.0249 | **3.19 (1.36 – 7.51)** | **0.0078** |
| Hypertension |  |  |  |  |  |  |
| No | 1 |  | 1 |  | 1 |  |
| Yes | 0.91 (0.48 – 1.72) | 0.7745 | 1.16 (0.61 – 2.20) | 0.6568 | 2.26 (0.96 – 5.29) | 0.0612 |
| ISI |  |  |  |  |  |  |
| ≤ 7 | 1 |  | 1 |  | 1 |  |
| > 7 | 0.44 (0.22 – 0.86) | 0.0157 | 0.53 (0.27 – 1.04) | 0.0659 | 1.39 (0.61 – 3.16) | 0.4375 |

The multinomial logistic regression analysis was performed on 1770 participants due to missing values on fast blood glucose levels. During the backward stepwise selection procedure, the following variables were not retained in the final model because they were not statistically significant: physical activity, alcohol use, tobacco use, PSQI, and diabetes. RLS-only was used as reference for the analysis. BMI, body mass index; ISI, insomnia severity index; OSA, obstructive sleep apnea; RLS, restless legs syndrome. COROSA, comorbid restless legs syndrome and obstructive sleep apnea; OR, odds-ratio; aOR, adjusted odds-ratio; CI, confidence interval. Model accuracy: 0.6378. Mean AUC: 0.756. AUC per class: 0.723 (None); 0.710 (OSA only); 0.754 (RLS only); 0.838 (COROSA). Mean AUC: 0.756.

### Exploratory analysis: hypertension burden across sleep-disorder phenotypes

In the exploratory analysis distinguishing eight mutually exclusive sleep-disorder phenotypes (Figure 1), hypertension was independently associated with both COROSA and COMIROSA (n = 27) compared to participants without any sleep disorder. The magnitude of association was higher for COMIROSA (aOR 4.03, 95% CI: 1.51–10.72) than for COROSA (aOR 2.64, 95% CI: 1.22–5.72). However, this difference was not statistically significant in a post hoc comparison of regression coefficients (p = 0.51).

**Fig. 1.**
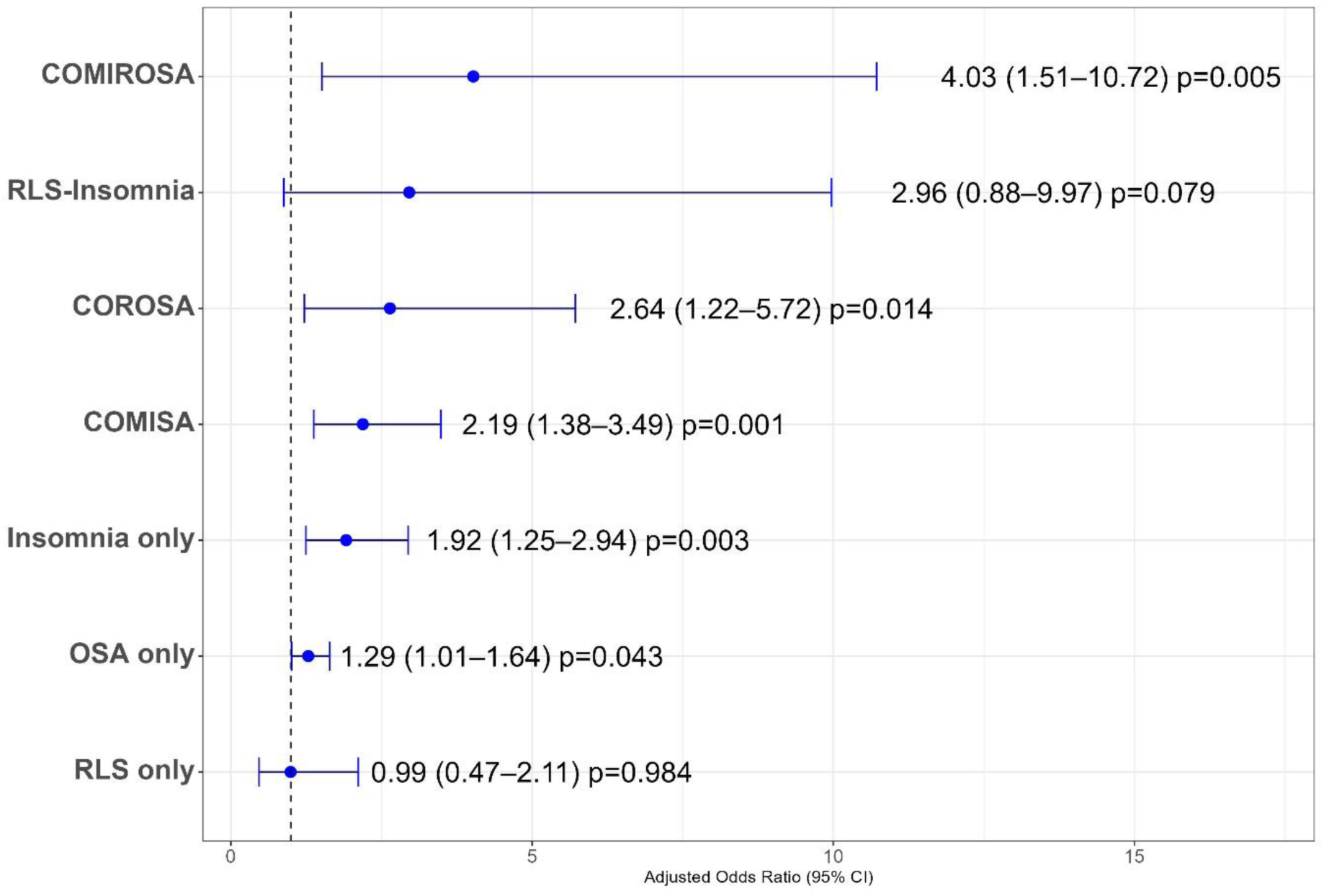
Sensitivity analysis: association of hypertension with eight mutually exclusive sleep-disorder phenotypes (multinomial logistic regression; N=1770) Points and horizontal bars represent adjusted odds ratios and 95% confidence intervals, respectively. The multinomial logistic regression model was adjusted for age, sex, area of residence, BMI, diabetes status, and sleep quality (PSQI). BMI, body mass index; OSA, obstructive sleep apnea; RLS, restless legs syndrome; PSQI, Pittsburgh Sleep Quality Index. COROSA; comorbid restless legs syndrome and obstructive sleep apnea; COMIROSA, comorbid insomnia, restless legs syndrome, and obstructive sleep apnea.

### Sensitivity analyses

The binary logistic regression analyses of factors associated with COROSA showed similar results when insomnia was defined using ISI thresholds of ≥ 10 or ≥ 15 (Supplementary Tables S2a and S2b). Likewise, the multinomial logistic regression analyses yielded comparable patterns of association (Supplementary Tables S3a and S3b). However, the exploratory analysis of hypertension across sleep-disorder phenotypes showed attenuation of the association with COMIROSA as more stringent insomnia thresholds were applied, with wider confidence intervals reflecting the smaller number of participants meeting these criteria. In contrast, the association between hypertension and COROSA remained statistically significant across insomnia definitions (Figures S1a & S1b).

## Discussion

This population-based study is, to our knowledge, the first to estimate the prevalence and determinants of comorbid obstructive sleep apnea and restless legs syndrome (COROSA) in an African setting.

The prevalence of COROSA was 3.5% (95% CI: 2.7–4.4), corresponding to roughly 1 in 28 individuals. COROSA was independently associated with older age, rural residence, obesity, hypertension, and insomnia. Multinomial analyses indicated that hypertension and insomnia were most strongly associated to COROSA compared with single-disorder categories. Although not highly prevalent at the community level, COROSA clustered among individuals with substantial cardiometabolic and sleep-related burden, underscoring its potential clinical and public health relevance in sub-Saharan Africa.

### Prevalence of COROSA

The prevalence of COROSA observed in our study was substantially lower than that reported in previous studies, mostly conducted in clinical settings that included only patients with diagnosed OSA, thereby enriching their samples with individuals more likely to present multiple sleep disorders. For instance, Kim et al., Stelzer et al., and Gothi et al. reported COROSA prevalences of 16.2%–24.4% among patients with OSA [4,7,21], and Pistorius et al. reported a prevalence of 20.7% in a clinical sample of sleep apnea syndrome (SAS) and RLS [20].

In contrast, our study was population-based, and included both symptomatic and asymptomatic individuals, providing a more representative estimate of the burden of COROSA in the general population. The lower prevalence observed in our study therefore likely reflects the underlying population burden rather than the higher estimates typically reported in the clinical cohorts subject to selection bias. This distinction is important for public health planning, as it suggests although COROSA is less frequent in the general population, it remains a non-negligible and clinically relevant condition.

### Factors associated with COROSA

The association between COROSA and older age is consistent with existing literature, as both OSA and RLS are known to increase with age [22–25]. Age-related physiological and neurological changes, including reduced upper-airway muscle tone and dopaminergic dysfunction [26], may be associated with a greater likelihood of OSA–RLS co-occurrence observed among older adults in our study.

The associations with overweight, obesity and hypertension further suggest an important cardiometabolic component. Obesity is a well-established risk factor for OSA and may also contribute to metabolic and inflammatory pathways implicated in RLS, whereas recurrent sleep disruption may, in turn, aggravate metabolic and cardiovascular dysfunction [27–31]. Hypertension has been reported more frequently in individuals with RLS and in those with coexisting OSA and RLS, who may exhibit heightened sympathetic activity and a greater burden of metabolic, psychiatric, and cognitive comorbidities [7,30–33]. Together, these findings suggest that COROSA may represent a more complex multisystem sleep-disorder profile rather than simply the coexistence of two independent conditions.

The strong association between COROSA and rural residence is noteworthy and has received limited attention in previous studies. In the same BeSAS population, rural residence was associated with higher odds of RLS, whereas urban residence was associated with substantially lower odds of the disorder (aOR 0.2, 95% CI 0.1–0.3) [12]. This disparity may partly reflect differences in nutritional status, including a potentially greater burden of iron deficiency in rural populations, given the central role of iron metabolism in RLS pathophysiology [34,35]. Differences in access to diagnosis and healthcare may also contribute. Although these mechanisms were not directly assessed, our findings underscore the need to consider nutritional, socioeconomic, and health-system factors when investigating COROSA in sub-Saharan Africa.

The multinomial analyses further clarified the phenotype of COROSA in relation to isolated sleep disorders. Older age was associated with OSA only, RLS only, and COROSA, although the associations were stronger for RLS only and COROSA, suggesting that aging may increase susceptibility to RLS-containing phenotypes. Male sex and overweight/obesity were more characteristic of OSA only, consistent with the established epidemiological profile of OSA, whereas rural residence and greater insomnia severity were more strongly associated with RLS only and COROSA.

Direct comparisons provided additional evidence that COROSA has a profile distinct from that of either disorder alone. Compared with isolated OSA, COROSA was associated with higher odds of hypertension and clinically significant insomnia, suggesting a greater combined cardiometabolic and sleep-related burden. However, the association with hypertension was no longer statistically significant when isolated RLS was used as the reference group, suggesting that the greater odds observed relative to isolated OSA may primarily reflect the RLS component of COROSA. By contrast, obesity was the principal factor distinguishing COROSA from isolated RLS, consistent with the role of excess body weight in the development of the OSA component of the overlap phenotype.

Insomnia emerged as a particularly important correlate of COROSA and is also a recurrent feature of OSA–RLS coexistence in previous studies. Clinical investigations by Stelzer et al. [21] and Pistorius et al. [20], reported a greater burden of insomnia symptoms among patients with coexisting OSA and RLS than among those with OSA alone. Our findings extend these observations to a community-based population and suggest that insomnia may be an integral feature of a more complex sleep disturbance profile involving both sleep-disordered breathing and impaired sleep continuity.

On this basis, we explored a broader overlap phenotype comprising insomnia, restless legs syndrome, and obstructive sleep apnea, which we tentatively termed COMIROSA. In the exploratory analysis, hypertension was associated with both COROSA and COMIROSA compared with the absence of sleep disorders, but the magnitude of the association was greater for COMIROSA than for COROSA (aOR 4.03 vs 2.64).

Although the analysis was not designed to compare these phenotypes directly and the number of COMIROSA cases was limited, this pattern raises the possibility that the addition of insomnia to coexisting RLS and OSA may confer an incremental cardiometabolic burden. Such an interpretation is biologically plausible because all three disorders have been linked to sleep fragmentation, sympathetic activation, daytime impairment, and adverse cardiovascular outcomes, and their coexistence may therefore produce additive or synergistic effects [36–38].

Exploratory sensitivity analyses suggested that the observed association between hypertension and COMIROSA depended on the operational definition of insomnia. Although the association was strongest when insomnia was defined using the conventional ISI threshold (>7), it attenuated with more stringent thresholds (ISI ≥ 10 and ≥ 15), likely reflecting the substantially smaller number of participants meeting the COMIROSA definition.

COMIROSA should nevertheless be regarded as a hypothesis-generating construct that requires formal definition and validation in larger epidemiological, clinical, and mechanistic studies.

### Clinical and public health implications

These findings have important implications for clinical practice and health policy in sub-Saharan Africa, where sleep disorders remain markedly under-recognized. In resource-limited settings, integrated and pragmatic screening strategies are needed. Prioritizing high-risk groups, older adults, individuals with obesity or hypertension, rural residents, and those reporting insomnia, may be a feasible approach to improving detection of COROSA. Clinicians evaluating patients with OSA should also consider systematic assessment for RLS and insomnia symptoms, while patients presenting with RLS or persistent insomnia may benefit from evaluation for underlying sleep-disordered breathing. These approaches could support earlier identification of multimorbid sleep conditions, although their feasibility and effectiveness in low-resource settings require further evaluation.

Future studies should determine whether integrated management of overlapping sleep disorders improves cardiometabolic and patient-reported outcomes compared with disorder-specific approaches.

### Strengths and limitations

This study has several strengths. Its population-based design and relatively large sample provided one of the first community-level estimates of COROSA in an African context and reduced referral and selection bias inherent in clinic-based cohorts. The use of standardized tools to assess OSA and RLS, along with multinomial modelling contrasting COROSA with OSA-only and RLS-only, allowed a nuanced characterization of risk profiles and supported the hypothesis of a distinct, more complex profile.

Some limitations should be acknowledged. The cross-sectional design precludes causal inference regarding the temporal relationships between COROSA and its associated factors. OSA was assessed using polygraphy rather than polysomnography, the diagnostic gold standard, which may have led to misclassification, especially of milder disease and sleep architecture disturbances that may be relevant to insomnia phenotyping. In addition, although the study was community-based, the sample may not fully reflect the diversity of the broader Beninese or sub-Saharan African populations, which may limit generalizability. Furthermore, the relatively small number of COROSA cases may have limited statistical precision for some estimates. Finally, the proposed COMIROSA concept was exploratory and not formally defined or validated as an outcome; its introduction here is conceptual and regarded as hypothesis-generating.

## Conclusion

In this population-based study, COROSA affected 3.5% of adults, providing the first community-based estimate in an African setting. Although less frequent than in clinical samples, it clustered among individuals with substantial cardiometabolic and sleep-related vulnerability. Older age, rural residence, overweight, obesity, hypertension, and insomnia were independently associated with COROSA, while secondary analyses suggested that it combines characteristics of both isolated OSA and isolated RLS. Together, these findings suggest that COROSA may represent a distinct, high-burden clinical profile rather than a simple coexistence of two sleep disorders. Our exploratory findings also raise the possibility of a more complex overlap involving insomnia, OSA and RLS (COMIROSA). Longitudinal, multicentre studies incorporating polysomnography and detailed phenotyping are needed to validate this proposed phenotype and determine its prognostic and therapeutic significance.

## Author contributions

Conceptualization: APW, GA, RH. Methodology: APW, RH, AS, HL, PMF, SS. Statistical analysis and interpretation: AS, APW. Writing – original draft preparation: AS, APW. Writing – review and editing: APW, AS, RH, EAD, AK, HL, PMF, SS, DM, DG, GA. Funding acquisition: RH, APW, GA. All authors reviewed and approved the final manuscript.

## Declarations

### Ethics approval

The research project was approved by the National Ethics Committee of Benin (reference number 45; Oct 25, 2017) with regular annual approval renewal throughout its course.

## Data Availability

Data are available from the corresponding author upon reasonable request.

## Acknowledgements

We acknowledge the contributions of the following persons who were involved in data acquisition on the field: Biaou Boni Richard, Adjiha Sylvia, Akpaki Ulrich, all the contributors on the field.

