## Supplementary material for "Prevalence and correlates of comorbid obstructive sleep apnea and restless legs syndrome in a population-based study from Benin, West Africa"

4. Exploratory analysis: hypertension burden across sleep-disorder phenotypes ... 9

1. **Distribution of patients' characteristics per sleep disorders phenotype**

Table S1: Baseline characteristics of BeSAS participants according to the sleep disorder status (N=1810)

| | None (n=981) | OSA only (n=718) | RLS only (n=48) | COROSA (n=63) | p-value |
|---|---|---|---|---|---|
| Age (years) | | | | | < 0.0001[a] |
| 25-39 | 539 (54.9%) | 210 (29.2%) | 9 (18.8%) | 9 (14.3%) | |
| 40-59 | 315 (32.1%) | 335 (46.7%) | 20 (41.7%) | 32 (50.8%) | |
| ≥60 | 127 (12.9%) | 173 (24.1%) | 19 (39.6%) | 22 (34.9%) | |
| Sex | | | | | < 0.0001[a] |
| Female | 679 (69.2%) | 404 (56.3%) | 35 (72.9%) | 45 (71.4%) | |
| Male | 302 (30.8%) | 314 (43.7%) | 13 (27.1%) | 18 (28.6%) | |
| Area of residence | | | | | < 0.0001[a] |
| Rural | 564 (57.5%) | 296 (41.2%) | 40 (83.3%) | 55 (87.3%) | |
| Urban | 417 (42.5%) | 422 (58.8%) | 8 (16.7%) | 8 (12.7%) | |
| BMI category | | | | | < 0.0001[a] |
| Underweight/Normal weight | 680 (69.3%) | 327 (45.5%) | 34 (70.8%) | 28 (44.4%) | |
| Overweight/Obese | 301 (30.7%) | 391 (54.5%) | 14 (29.2%) | 35 (55.6%) | |
| Physical activity | | | | | 0.4733[a] |
| No | 803 (81.9%) | 570 (79.4%) | 38 (79.2%) | 48 (76.2%) | |
| Yes | 178 (18.1%) | 148 (20.6%) | 10 (20.8%) | 15 (23.8%) | |
| Alcohol use | | | | | 0.0039[a] |
| No | 896 (91.3%) | 620 (86.4%) | 42 (87.5%) | 52 (82.5%) | |
| Yes | 85 (8.7%) | 98 (13.3%) | 28 (12.5%) | 11 (17.5%) | |
| Tobacco use | | | | | < 0.0001[a] |
| No | 920 (93.8%) | 654 (91.1%) | 38 (79.2%) | 52 (82.5%) | |
| Yes | 61 (6.2%) | 64 (8.9%) | 10 (20.8%) | 11 (17.5%) | |
| Hypertension | | | | | < 0.0001[a] |
| No | 614 (62.6%) | 320 (44.6%) | 23 (47.9%) | 17 (27.0%) | |
| Yes | 367 (37.4%) | 398 (55.4%) | 25 (52.1%) | 15 (73.0%) | |
| Diabetes* | | | | | 0.0008[b] |

| | | | | | |
|---|---|---|---|---|---|
| No | 947 (97.4%) | 644 (93.6%) | 47 (97.9%) | 58 (93.5%) | |
| Yes | 25 (2.6%) | 44 (6.4%) | 1 (2.1%) | 4 (6.5%) | |
| PSQI | | | | | 0.0023[a] |
| < 6 | 468 (47.7%) | 330 (46.0%) | 15 (31.2%) | 17 (27.0%) | |
| ≥ 6 | 513 (52.3%) | 388 (54.0%) | 33 (68.8%) | 46 (73.0%) | |
| ISI | | | | | < 0.0001[a] |
| ≤ 7 | 863 (88.0%) | 600 (83.6%) | 33 (68.8%) | 36 (57.1%) | |
| > 7 | 118 (12.0%) | 118 (16.4%) | 15 (31.2%) | 27 (42.9%) | |
| ESS | | | | | 0.3871[a] |
| < 11 | 831 (84.7%) | 587 (81.8%) | 39 (81.2%) | 51 (81.0%) | |
| ≥ 11 | 150 (15.3%) | 131 (18.2%) | 9 (18.8%) | 12 (19.0%) | |

*Data on diabetes were analyzed on a total sample of 1770 participants due to missing values on fast blood glucose levels. BMI, body mass index; PSQI, Pittsburgh Sleep Quality Index; ESS, Epworth Sleepiness Scale. ISI, insomnia severity index; OSA, obstructive sleep apnea; RLS, restless legs syndrome. COROSA, comorbid restless legs syndrome and obstructive sleep apnea. [a]Pearson's chi-squared test; [b]Fisher's exact test.

### 2. Factors associated with COROSA

Table S2a: Factors associated with COROSA among BeSAS participants – binary logistic regression analysis using a 10 ISI score threshold (N=1810)

| | aOR | 95% CI | p-value |
|---|---|---|---|
| Age (years) | | | |
| 25-39 | 1 | | |
| 40-59 | 3.01 | 1.43 – 6.94 | 0.0057 |
| ≥60 | 3.89 | 1.67 – 9.66 | 0.0022 |
| Sex | | | |
| Female | 1 | | |
| Male | 0.88 | 0.45 – 1.67 | 0.7132 |
| Area of residence | | | |
| Urban | 1 | | |
| Rural | 10.67 | 4.99 – 25.75 | < 0.0001 |
| BMI category | | | |
| Normal weight | 1 | | |
| Underweight | 1.29 | 0.51 – 3.00 | 0.5628 |
| Overweight | 2.29 | 1.17 – 4.50 | 0.0150 |
| Obese | 3.95 | 1.81 – 8.55 | 0.0004 |
| Alcohol use | | | |
| No | 1 | | |
| Yes | 1.37 | 0.60 – 2.93 | 0.4302 |
| Tobacco use | | | |
| No | 1 | | |
| Yes | 1.39 | 0.61 – 2.98 | 0.4139 |
| Hypertension | | | |
| No | 1 | | |
| Yes | 2.41 | 1.33 – 4.54 | 0.0045 |
| PSQI | | | |
| < 6 | 1 | | |
| ≥ 6 | 1.32 | 0.72 – 2.50 | 0.3850 |
| ISI | | | |
| < 10 | 1 | | |
| ≥ 10 | 2.19 | 1.14 – 4.09 | 0.0154 |

The multivariable model was adjusted for age, sex, area of residence, BMI category, alcohol use, tobacco use, hypertension, PSQI and insomnia severity. aOR, adjusted odds-ratio; CI, confidence interval; BMI, body mass index; PSQI, Pittsburgh Sleep Quality Index; ISI, insomnia severity index. Model performance – Hosmer-Lemeshow Goodness-of-Fit Test: p-value = 0.862; AUC: 0.842 (95% CI: 0.793 – 0.891).

Table S2b: Factors associated with COROSA among BeSAS participants – binary logistic regression analysis using a 15 ISI score threshold (N=1810)

| | **aOR** | **95% CI** | **p-value** |
|---|---|---|---|
| Age (years) | | | |
| 25-39 | 1 | | |
| 40-59 | 3.00 | 1.42 – 6.92 | 0.0059 |
| ≥60 | 3.79 | 1.64 – 9.39 | 0.0025 |
| Sex | | | |
| Female | 1 | | |
| Male | 0.86 | 0.44 – 1.64 | 0.6565 |
| Area of residence | | | |
| Urban | 1 | | |
| Rural | 10.75 | 5.02 – 26.05 | < 0.0001 |
| BMI category | | | |
| Normal weight | 1 | | |
| Underweight | 1.29 | 0.50 – 3.00 | 0.5765 |
| Overweight | 2.36 | 1.20 – 4.65 | 0.0126 |
| Obese | 4.14 | 1.88 – 9.07 | 0.0004 |
| Alcohol use | | | |
| No | 1 | | |
| Yes | 1.45 | 0.64 – 3.09 | 0.3522 |
| Tobacco use | | | |
| No | 1 | | |
| Yes | 1.48 | 0.65 – 3.15 | 0.3303 |
| Hypertension | | | |
| No | 1 | | |
| Yes | 2.58 | 1.42 – 4.88 | 0.0024 |
| PSQI | | | |
| < 6 | 1 | | |
| ≥ 6 | 1.32 | 0.72 – 2.48 | 0.3803 |
| ISI | | | |
| < 15 | 1 | | |
| ≥ 15 | 4.93 | 2.07 – 11.06 | 0.0002 |

The multivariable model was adjusted for age, sex, area of residence, BMI category, alcohol use, tobacco use, hypertension, PSQI and insomnia severity. cOR, crude odds-ratio; aOR, adjusted odds-ratio; CI, confidence interval; BMI, body mass index; PSQI, Pittsburgh Sleep Quality Index; ISI, insomnia severity index. Model performance – Hosmer-Lemeshow Goodness-of-Fit Test: p-value = 0.938; AUC: 0.841 (95% CI: 0.791 – 0.892).

3. **Comparative analysis of determinants of OSA-only, RLS-only and COROSA**

Table S3a: Multivariable analyses of factors associated with COROSA among BeSAS participants – multinomial logistic regression using a 10 ISI score threshold (N=1770)

| | **OSA only** | | **RLS only** | | **COROSA** | |
|---|---|---|---|---|---|---|
| | **aOR (95% CI)** | **p-value** | **aOR (95% CI)** | **p-value** | **aOR (95% CI)** | **p-value** |
| Age (years) | | | | | | |
| 25-39 | 1 | | 1 | | 1 | |
| 40-59 | 2.29 (1.80 – 2.91) | < 0.0001 | 3.50 (1.54 – 8.00) | 0.0028 | 4.47 (2.04 – 9.81) | 0.0002 |
| ≥60 | 3.16 (2.31 – 4.31) | < 0.0001 | 7.67 (3.21 – 18.3) | < 0.0001 | 7.59 (3.22 – 17.87) | < 0.0001 |
| Sex | | | | | | |
| Female | 1 | | 1 | | 1 | |
| Male | 2.39 (1.90 – 3.02) | < 0.0001 | 0.95 (0.48 – 1.88) | 0.8811 | 1.50 (0.82 – 2.77) | 0.1915 |
| Area of residence | | | | | | |
| Urban | 1 | | 1 | | 1 | |
| Rural | 0.78 (0.63 – 0.97) | 0.0285 | 4.11 (1.85 – 9.17) | 0.0005 | 10.13 (4.43–23.18) | < 0.0001 |
| BMI category | | | | | | |
| Underweight/Normal weight | 1 | | 1 | | 1 | |
| Overweight/Obese | 2.90 (2.29 – 3.68) | < 0.0001 | 1.32 (0.66 – 2.63) | 0.4337 | 4.26 (2.38 – 7.62) | < 0.0001 |
| Hypertension | | | | | | |
| No | 1 | | 1 | | 1 | |
| Yes | 1.28 (1.02 – 1.61) | 0.0325 | 1.13 (0.60 – 2.12) | 0.7095 | 2.68 (1.44 – 4.98) | 0.0017 |
| ISI | | | | | | |
| < 10 | 1 | | 1 | | 1 | |
| ≥ 10 | 1.18 (0.81 – 1.71) | 0.3923 | 3.08 (1.51 – 6.28) | 0.0019 | 2.87 (1.51 – 5.47) | 0.0013 |

The multinomial logistic regression analysis was performed on 1770 participants due to missing values on fast blood glucose levels. During the backward stepwise selection procedure, the following variables were not retained in the final model because they were not statistically significant: physical activity, alcohol use, tobacco use, PSQI, and diabetes. “No OSA and no RLS” was used as reference for the analysis. BMI, body mass index; ISI, insomnia severity index; OSA, obstructive sleep apnea; RLS, restless legs syndrome.

COROSA, comorbid restless legs syndrome and obstructive sleep apnea; OR, odds-ratio; aOR, adjusted odds-ratio; CI, confidence interval. Model accuracy: 0.6355. AUC per class: 0.723 (None); 0.710 (OSA only); 0.761 (RLS only); 0.836 (COROSA). Mean AUC: 0.757.

Table S3b: Multivariable analyses of factors associated with COROSA among BeSAS participants – multinomial logistic regression using a 15 ISI score threshold (N=1770)

| | OSA only | | RLS only | | COROSA | |
|---|---|---|---|---|---|---|
| | **aOR (95% CI)** | **p-value** | **aOR (95% CI)** | **p-value** | **aOR (95% CI)** | **p-value** |
| Age (years) | | | | | | |
| 25-39 | 1 | | 1 | | 1 | |
| 40-59 | 2.31 (1.81 – 2.94) | < 0.0001 | 3.71 (1.64 – 8.43) | 0.0017 | 4.57 (2.08 – 10.02) | 0.0001 |
| ≥60 | 3.18 (2.33 – 4.34) | < 0.0001 | 7.99 (3.35 – 19.06) | < 0.0001 | 7.51 (3.18 – 17.72) | < 0.0001 |
| Sex | | | | | | |
| Female | 1 | | 1 | | 1 | |
| Male | 2.37 (1.89 – 2.99) | < 0.0001 | 0.91 (0.46 – 1.80) | 0.7877 | 1.49 (0.80 – 2.75) | 0.2074 |
| Area of residence | | | | | | |
| Urban | 1 | | 1 | | 1 | |
| Rural | 0.78 (0.63 – 0.97) | 0.0281 | 4.08 (1.82 – 9.15) | 0.0006 | 10.25 (4.46–23.55) | < 0.0001 |
| BMI category | | | | | | |
| Underweight/Normal weight | 1 | | 1 | | 1 | |
| Overweight/Obese | 2.90 (2.29 – 3.67) | < 0.0001 | 1.33 (0.66 – 2.67) | 0.4199 | 4.39 (2.44 – 7.89) | < 0.0001 |
| Hypertension | | | | | | |
| No | 1 | | 1 | | 1 | |
| Yes | 1.29 (1.03 – 1.61) | 0.0296 | 1.20 (0.64 – 2.27) | 0.5638 | 2.94 (1.58 – 5.47) | 0.0007 |
| ISI | | | | | | |
| < 15 | 1 | | 1 | | 1 | |
| ≥ 15 | 1.17 (0.54 – 2.53) | 0.6837 | 5.08 (1.81 – 14.24) | 0.0020 | 6.71 (2.67 – 16.85) | < 0.0001 |

The multinomial logistic regression analysis was performed on 1770 participants due to missing values on fast blood glucose levels. During the backward stepwise selection procedure, the following variables were not retained in the final model because they were not statistically significant: physical activity, alcohol use, tobacco use, PSQI, and diabetes. “No OSA and no RLS” was used as reference for the analysis. BMI, body mass index; ISI, insomnia severity index; OSA, obstructive sleep apnea; RLS, restless legs syndrome. COROSA, comorbid restless legs syndrome and obstructive sleep apnea; OR, odds-ratio; aOR, adjusted odds-ratio; CI, confidence interval. Model accuracy: 0.6378. AUC per class: 0.723 (None); 0.710 (OSA only); 0.754 (RLS only); 0.838 (COROSA). Mean AUC: 0.756.

### 4. Exploratory analysis: hypertension burden across sleep-disorder phenotypes

- ISI threshold of ≥ 10

| None (n=900) | OSA only (n=619) | RLS only (n=35) | Insomnia only (n=72) | COMISA (n=69) | RLS-Insomnia (n=13) | COROSA (n=44) | COMIROSA (n=18) |
|---|---|---|---|---|---|---|---|

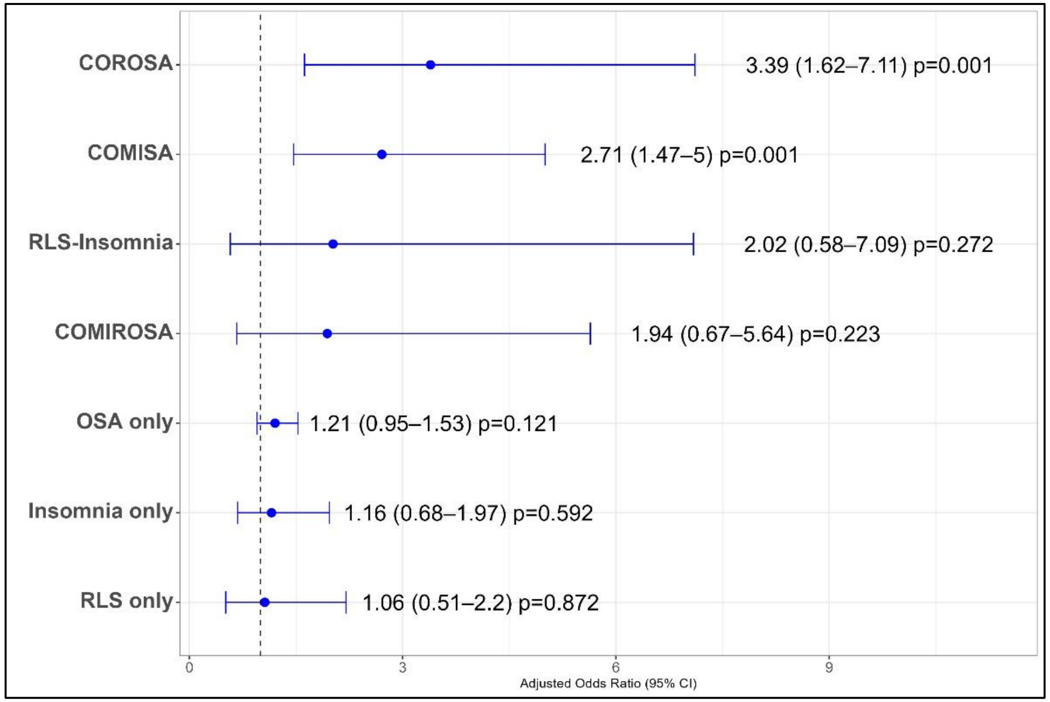

Figure S1a: Sensitivity analysis: association of hypertension with eight mutually exclusive sleep-disorder phenotypes using ISI threshold of ≥ 10 (multinomial logistic regression; N=1770)

Points and horizontal bars represent adjusted odds ratios and 95% confidence intervals, respectively. The multinomial logistic regression model was adjusted for age, sex, area of residence, BMI, diabetes status, and sleep quality (PSQI). BMI, body mass index;

OSA, obstructive sleep apnea; RLS, restless legs syndrome; PSQI, Pittsburgh Sleep Quality Index; ISI, insomnia severity index COROSA; comorbid restless legs syndrome and obstructive sleep apnea; COMIROSA, comorbid insomnia, restless legs syndrome, and obstructive sleep apnea.

- ISI threshold of ≥ 15

| None (n=957) | OSA only (n=673) | RLS only (n=42) | Insomnia only (n=15) | COMISA (n=15) | RLS-Insomnia (n=6) | COROSA (n=52) | COMIROSA (n=10) |
|---|---|---|---|---|---|---|---|

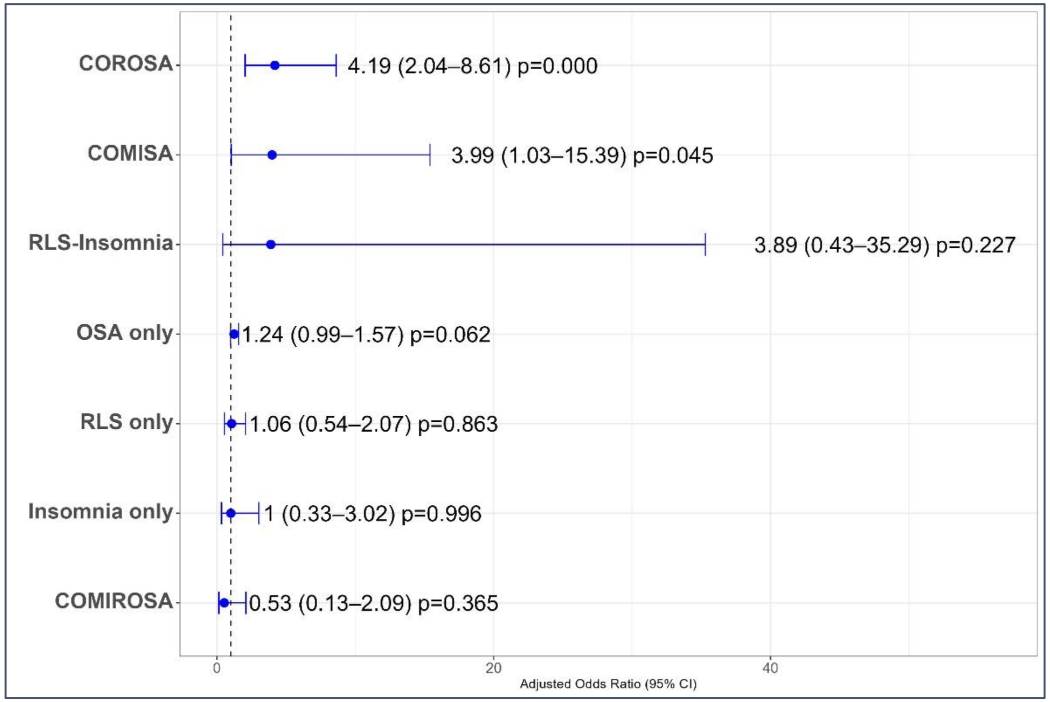

Figure S1b: Sensitivity analysis: association of hypertension with eight mutually exclusive sleep-disorder phenotypes ISI threshold of ≥ 15 (multinomial logistic regression; N=1770)

Points and horizontal bars represent adjusted odds ratios and 95% confidence intervals, respectively. The multinomial logistic regression model was adjusted for age, sex, area of residence, BMI, diabetes status, and sleep quality (PSQI). BMI, body mass index; OSA, obstructive sleep apnea; RLS, restless legs syndrome; PSQI, Pittsburgh Sleep Quality Index; ISI, insomnia severity index; COROSA, comorbid restless legs syndrome and obstructive sleep apnea; COMIROSA, comorbid insomnia, restless legs syndrome, and obstructive sleep apnea.
